# A Deep Learning–Based Automated Detection of Mucus Plugs in Chest CT

**DOI:** 10.1101/2025.03.09.25323501

**Authors:** Yuki Sonoda, Kensuke Fukuda, Hirotaka Matsuzaki, Yosuke Yamagishi, Soichiro Miki, Yukihiro Nomura, Yu Mikami, Takeharu Yoshikawa, Shouhei Hanaoka, Hidenori Kage, Osamu Abe

## Abstract

This study presents a novel two stage deep learning algorithm for automated detection of mucus plugs in CT scans of patients with respiratory diseases. Despite the clinical significance of mucus plugs in COPD and asthma where they indicate hypoxemia, reduced exercise tolerance, and poorer outcomes, they remain under evaluated in clinical practice due to labor intensive manual annotation. The developed algorithm first segments both patent and obstructed airways using a VNet-based model pre-trained on normal airway structures and fine-tuned on mucus containing scans. Subsequently, a rule-based post processing method identifies mucus plugs by evaluating cross sectional areas along airway centerlines. Validation on an in-house dataset of 33 CT scans from patients with asthma/COPD demonstrated high sensitivity (93.8%) though modest positive predictive value (18.8%). Performance on an external dataset (LIDC-IDRI) achieved 82.8% sensitivity with 23.5% PPV. While challenges remain in reducing false positives, this automated detection tool shows promise for screening applications in both clinical and research settings, potentially addressing the current gap in mucus plug evaluation within standard practice.

## Introduction

In COPD the airway mucus plugs observed in chest CT indicate hypoxemia independently of emphysema and are associated with exercise tolerance, exacerbation frequency, and life expectancy.^1^ In asthma, they are further associated with lung function, severity, and treatment response.^2^ Dunican et al. defined mucus plugs as complete occlusion of a bronchus, irrespective of generation or size, excluding the most peripheral bronchi within 2 cm of the costal pleura or diaphragmatic surface.^2^ Although CT results of only a small proportion of healthy controls (approximately 5–10%) present mucus plugs, they may be present in a considerable proportion (approximately 50%) in patients with severe asthma or COPD.^3^

Despite their clinical importance, mucus plugs are rarely evaluated in standard clinical practice owing to labor-intensive manual annotation. To our knowledge, no automated detection programs have been validated against the gold-standard method of visual detection based on Dunican et al.’s definition.^4^ Furthermore, despite advances in deep learning techniques in the medical domain, factors such as bronchial wall thickening and partial volume effects challenge the elucidation of the ground truth. Dournes et al. reported a deep learning-based detection method for airway lesions including mucus plugs in patients with cystic fibrosis,^5^ where such plugs are abundant. However, many patients with COPD or asthma present with very few or no plugs, making their detection difficult when using conventional deep-learning approaches, especially with limited training data.

The aim of this study was to develop an algorithm capable of detecting mucus plugs with high sensitivity. Rather than directly labeling voxels as “mucus”, we present a two-stage algorithm which first segments the entire airway including possible wall thickening and intraluminal material, and then identifies regions that are likely occluded by mucus.

## Methods

A VNet-based^6^ deep-learning model was pre-trained to segment normal airway structures using the Multi-site, Multi-domain Airway Tree Modeling (ATM’22) dataset.^7^ Subsequently, the model was fine-tuned using 10 high-resolution CT scans (in-plane resolution: 0.38–0.45 mm; slice thickness: 1.0 mm) from patients who had mucus noted in their radiology reports at our institution. A radiology resident with 3 years of experience labeled both the patent airway lumen and any hyperdense intraluminal area as a single structure. The model was trained on this unified label to segment both patent and obstructed airways.

Based on the model’s segmentation mask, a rule-based post-processing was applied to detect actual mucus plugs. High- and low-attenuation regions were distinguished using the method described by Yen et al.^8^ The airway centerlines were skeletonized, and each cross-section at centerline voxels was evaluated. If high-attenuation voxels occupied >95% of the cross-sectional area, the region was labeled a plug, a threshold selected to reduce false negatives caused by partial volume effects. Additionally, isolated clusters <1 µL were removed to reduce noise. Finally, an exclusion mask was generated using a whole-lung mask, a mediastinal mask using TotalSegmentator,^9^ and a mediastinal segmentation network trained using the ATM’22 dataset, separating peripheral airway branches. The trained weights and details of the code are openly available at https://github.com/miscelllaneous/MucusPlugDetector.

For validation, 33 in-house high-resolution CT scans (in-plane resolution: 0.39-0.47 mm; slice thickness: 1.0 mm) were selected from patients with asthma and/or COPD, who had no pneumonia or lung mass. Additionally, we extracted scans with an in-plane resolution of ≤0.55 mm and a slice thickness of ≤1 mm from the Lung Image Database Consortium-Image Database Resource Initiative (LIDC-IDRI)^10^ and excluded images that were part of the ATM’22 dataset, resulting in 14 eligible cases (in-plane resolution: 0.46-0.55 mm; slice thickness: 0.5-1.0 mm). Information regarding comorbidities, including COPD or asthma, was not available for the cases. A pulmonologist with 12 years of experience and the same radiology resident annotated and confirmed mucus plugs in each scan. Disagreements were resolved through consensus discussion. A detected plug was defined as any predicted region overlapping a ground-truth plug. Conversely, any predicted region not overlapping with the ground-truth annotation was considered a false positive. Subsequently, sensitivity and positive predictive value (PPV) were calculated for both datasets.

## Results

An overview of the results is presented in Figure 1.

**Figure 1.**
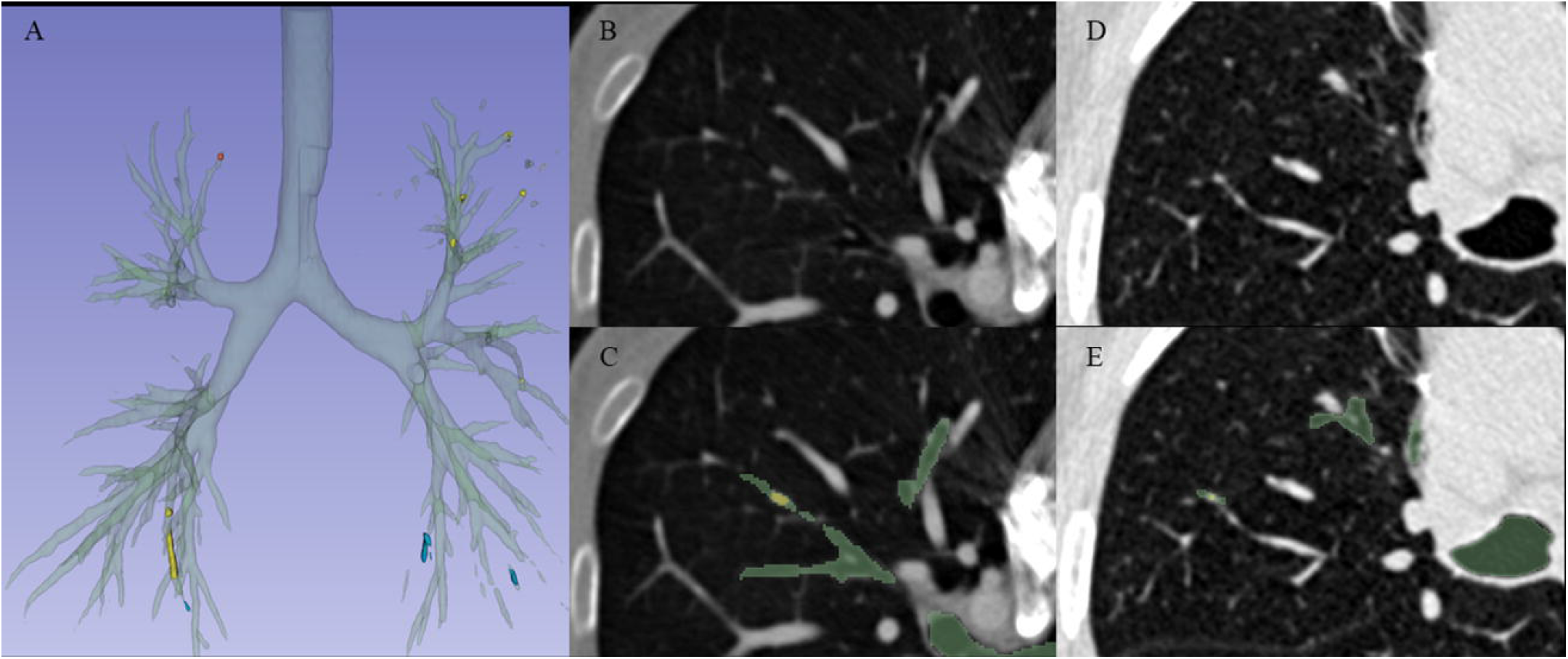
Overview of the algorithm output. (A) Frontal view of airway and mucus plug predictions with three-dimensional reconstruction. (B and D) Axial views showing examples of detected mucus plugs and (C and E) their corresponding segmentation masks. Green, areas classified as airways or mucus; Yellow, true-positives; Blue, false-positives; and Red, false-negatives.

In the 33 in-house and 14 LIDC-IDRI CT scans, mucus plugs ranged from 0–74 and 0–17, respectively (Table 1). The developed algorithm achieved 93.8% (228/243) sensitivity in the in-house dataset, with a PPV of 18.8% (228/1211). In contrast, the algorithm achieved a sensitivity of 82.8% (24/29) in the external LIDC-IDRI dataset, with a PPV of 23.5% (24/102). False positives were primarily attributed to misclassification of thickened bronchi, partial volume effects, motion artifacts, or adjacent vessels as obstructed airways.□False positives were particularly common in cases with poor breath-holding or motion artifacts.

**Table 1.**
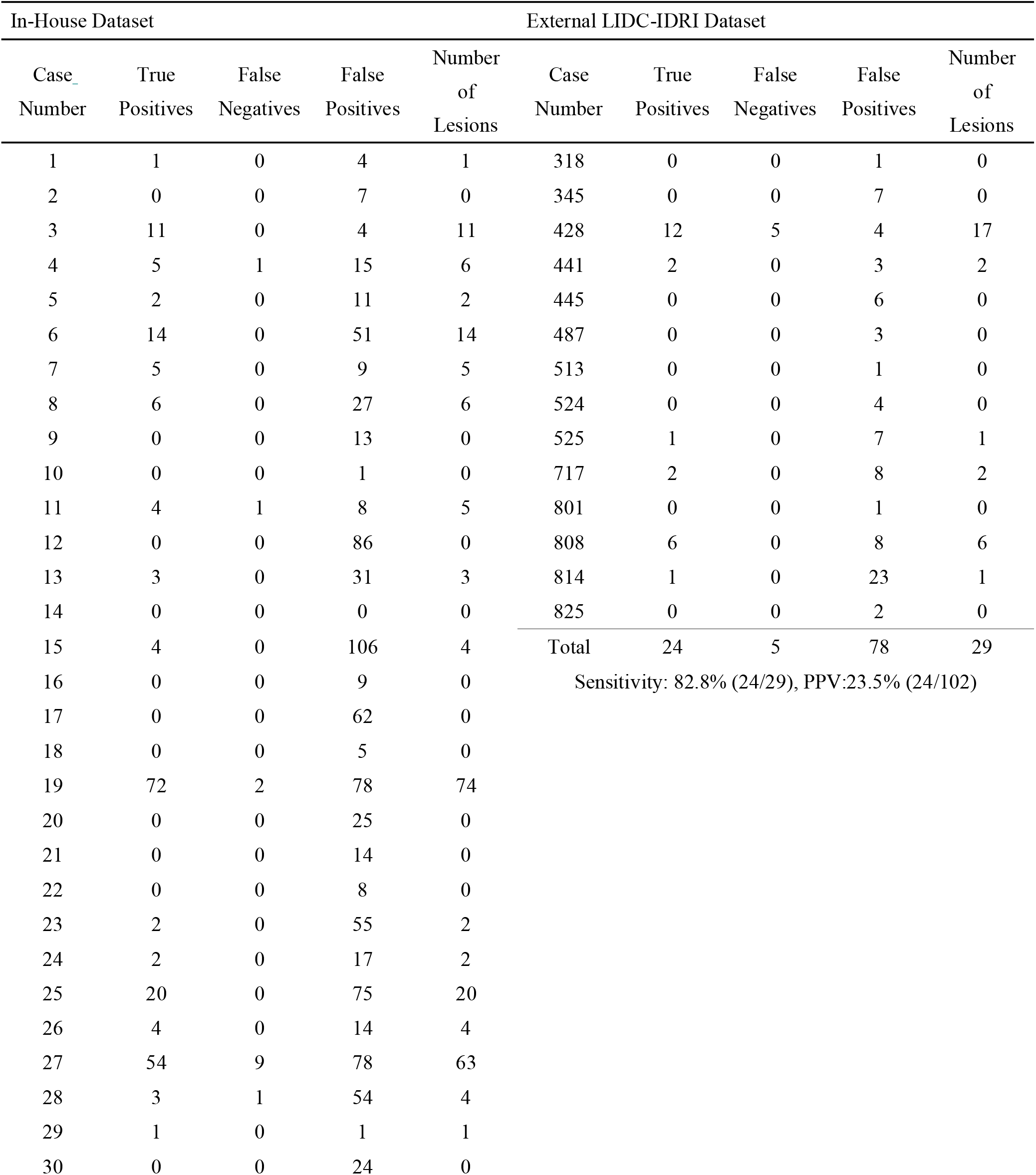
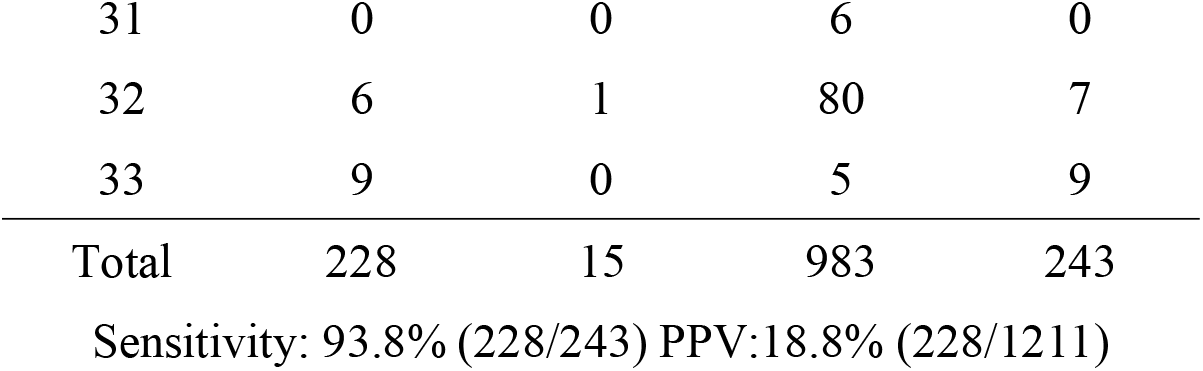
Per-case performance for the in-house and external Lung Image Database Consortium-Image Database Resource Initiative datasets. LIDC, Lung Image Database Consortium; IDRI, Image Database Resource Initiative; PPV, Positive Predictive Value.

| In-House Dataset |  |  |  |  | External LIDC-IDRI Dataset |  |  |  |  |
| --- | --- | --- | --- | --- | --- | --- | --- | --- | --- |
| Case_<br>Number | True<br>Positives | False<br>Negatives | False<br>Positives | Number<br>of<br>Lesions | Case<br>Number | True<br>Positives | False<br>Negatives | False<br>Positives | Number<br>of<br>Lesions |
| 1 | 1 | 0 | 4 | 1 | 318 | 0 | 0 | 1 | 0 |
| 2 | 0 | 0 | 7 | 0 | 345 | 0 | 0 | 7 | 0 |
| 3 | 11 | 0 | 4 | 11 | 428 | 12 | 5 | 4 | 17 |
| 4 | 5 | 1 | 15 | 6 | 441 | 2 | 0 | 3 | 2 |
| 5 | 2 | 0 | 11 | 2 | 445 | 0 | 0 | 6 | 0 |
| 6 | 14 | 0 | 51 | 14 | 487 | 0 | 0 | 3 | 0 |
| 7 | 5 | 0 | 9 | 5 | 513 | 0 | 0 | 1 | 0 |
| 8 | 6 | 0 | 27 | 6 | 524 | 0 | 0 | 4 | 0 |
| 9 | 0 | 0 | 13 | 0 | 525 | 1 | 0 | 7 | 1 |
| 10 | 0 | 0 | 1 | 0 | 717 | 2 | 0 | 8 | 2 |
| 11 | 4 | 1 | 8 | 5 | 801 | 0 | 0 | 1 | 0 |
| 12 | 0 | 0 | 86 | 0 | 808 | 6 | 0 | 8 | 6 |
| 13 | 3 | 0 | 31 | 3 | 814 | 1 | 0 | 23 | 1 |
| 14 | 0 | 0 | 0 | 0 | 825 | 0 | 0 | 2 | 0 |
| 15 | 4 | 0 | 106 | 4 | Total | 24 | 5 | 78 | 29 |
| 16 | 0 | 0 | 9 | 0 | Sensitivity: 82.8% (24/29), PPV:23.5% (24/102) |  |  |  |  |
| 17 | 0 | 0 | 62 | 0 |  |  |  |  |  |
| 18 | 0 | 0 | 5 | 0 |  |  |  |  |  |
| 19 | 72 | 2 | 78 | 74 |  |  |  |  |  |
| 20 | 0 | 0 | 25 | 0 |  |  |  |  |  |
| 21 | 0 | 0 | 14 | 0 |  |  |  |  |  |
| 22 | 0 | 0 | 8 | 0 |  |  |  |  |  |
| 23 | 2 | 0 | 55 | 2 |  |  |  |  |  |
| 24 | 2 | 0 | 17 | 2 |  |  |  |  |  |
| 25 | 20 | 0 | 75 | 20 |  |  |  |  |  |
| 26 | 4 | 0 | 14 | 4 |  |  |  |  |  |
| 27 | 54 | 9 | 78 | 63 |  |  |  |  |  |
| 28 | 3 | 1 | 54 | 4 |  |  |  |  |  |
| 29 | 1 | 0 | 1 | 1 |  |  |  |  |  |
| 30 | 0 | 0 | 24 | 0 |  |  |  |  |  |

|  |  |  |  |  |
| --- | --- | --- | --- | --- |
| 31 | 0 | 0 | 6 | 0 |
| 32 | 6 | 1 | 80 | 7 |
| 33 | 9 | 0 | 5 | 9 |
| Total | 228 | 15 | 983 | 243 |

Sensitivity: 93.8% (228/243) PPV:18.8% (228/1211)

## Discussion

The two-stage deep learning-based algorithm developed in this study, which first segments both patent and mucus-filled airways and then uses a rule-based method to detect mucus, achieved high sensitivity in mucus plug detection using the in-house dataset of patients with asthma or COPD, while maintaining a reasonable performance on the external dataset despite differences in imaging parameters and background of patients. To the best of our knowledge, no previous study has reported an automated tool for mucus plug detection, which has been validated against human visual standards.

In the external dataset, the sensitivity was inferior to our in-house results. Possible factors for the lower performance include the relatively coarser in-plane resolution, the inclusion of scans with suboptimal breath-holding technique, and the limitation that our airway extraction model was trained exclusively on in-house data. These performance constraints could potentially be addressed by enriching training data with diverse resolutions and expanding the variety of data sources.

Several challenges remain, including reducing the high false-positive rate and the presence of the rule-based component, which could potentially limit detection performance across different scanning protocols and resolutions. Nevertheless, the robust sensitivity and adaptability to external data demonstrated by the developed system suggests its efficacy as a screening tool, which can ease workloads in both clinical and research settings. Furthermore, the training data obtained through this approach, combined with deep-learning technology, may provide insights for the development of a fully automated tool with enhanced sensitivity and PPV.

## Data Availability

All data produced in the present study may be available upon reasonable request to the authors, with the exception of patient imaging data. Due to personal information protection considerations, the patient image data cannot be made publicly available in order to protect patient privacy and comply with relevant data protection regulations.

## Abbreviations

ATM’22

Multi-site: Multi-domain Airway Tree Modeling
COPD: chronic obstructive pulmonary disease
CT: computed tomography
IDRI: Image Database Resource Initiative
LIDC: Lung Image Database Consortium
PPV: positive predictive value.

